# Dynamic evaluation of spine kinematics in individuals with Parkinson’s disease and freezing of gait

**DOI:** 10.1101/2023.10.18.23297195

**Authors:** C. Palmisano, V. Farinelli, F. Camuncoli, A. Favata, G. Pezzoli, C.A. Frigo, I.U. Isaias

## Abstract

**Background:** Freezing of gait (FoG) is an episodic failure of gait exposing people with Parkinson’s disease (PD) to a high risk of falling. Despite growing evidence of the interconnection between impaired trunk control and FoG, a detailed description of spinal kinematics during walking is still lacking in this population.

**Research question:** Do spinal alterations impact gait performance in individuals with PD and FoG?

**Methods:** We analyzed kinematic data of 47 PD participants suffering (PD-FOG, N=24) or not suffering from FoG (PD-NFOG, N=23) and 15 healthy controls (HCO) during quiet standing and unperturbed walking. We estimated the main spinal variables (i.e., spinal length, lordosis and kyphosis angles, trunk inclination), the pelvis angles, and the shoulder-pelvis angles during gait and standing. We studied differences across conditions and groups and the relationships between postural and gait parameters using linear regression methods.

**Results:** During standing and walking, both PD groups showed increased trunk inclination and decreased lordosis angle with respect to HCO, as well as a decreased range in variation of kyphosis angle, pelvic obliquity, and shoulder-pelvis angles. Only PD-FOG participants showed reduced range of lordosis angle and spinal length compared to HCO. PD-FOG individuals were also not able to straighten their spine during walking compared to standing. Stride length and velocity were decreased in both patient groups compared to HCO, while swing duration was reduced only in the PD-FOG group. In individuals with PD-FOG, trunk inclination and lordosis angle showed moderate but significant positive correlations with all gait alterations.

**Significance:** Spine alterations impacted gait performance in individuals with PD suffering from FoG. Excessive trunk inclination and poor mastering of the lordosis spinal region may create an unfavourable postural precondition for forward walking. Physical therapy should target combined spinal and stepping alterations in these individuals.

## Introduction

Freezing of gait (FoG) is a severe gait disturbance affecting individuals with Parkinson’s disease (PD), causing a sudden inability to step forward despite the intention to walk [1]. Gait freezing often leads to falls [2,3], which contribute to injuries [4–8], fractures [6–8], fear of falling [4–7,9] and, in turn, poor quality [7,10] and quantity of life [8,9,11]. Investigating kinematic alterations underlying gait freezing and risk of falling is thus of utmost importance to improve fall prediction and prevention in PD.

Growing evidence indicates that alterations in the postural framework and gait derangements are two interconnected phenomena [4]. Gait in PD is typically characterized by forward trunk flexion [12–15], particularly in subjects prone to falls, and possibly connected to spinal muscular deficits [16]. A reduction in trunk range of motion (RoM) has also been described [17]. Of relevance, while dopaminergic medication and deep brain stimulation were shown to normalize trunk inclination [13,14], improvements in trunk stiffness were variable [18]. During gait freezing, patients report to feel their feet “glued to the floor”, while the trunk keeps moving forward [1]. Contributing to approximately 50% of the body’s mass, the trunk plays a fundamental role in balance control, particularly during dynamic motor tasks [16,19]. Misalignment between the lower limbs and the trunk may lead to an imbalance in the centre of mass (CoM), favouring forward falls [4]. In line, FoG is associated with falling forward while other clinical features of PD, such as impaired balance, akinetic-rigid subtype and neuropsychiatric symptoms, are related to falling in multiple directions [20].

Although previous studies and clinical evidence clearly indicate the interconnection between postural alterations and postural instability in FoG [4], research into the impact of spine kinematics on posture and gait has been mainly limited to healthy people. Trunk flexion during walking caused a crouch posture characterized by prolonged knee flexion and increased ankle dorsiflexion and hip flexion [21]. These postural adaptations resemble the typical parkinsonian postural asset [22,23], and may be the expression of compensatory strategies to counteract the anterior shift in the CoM caused by increased trunk flexion [21]. These postural changes may decrease the risk of falling forward, but they lead to a backward shift in the centre of pressure (CoP) during upright posture and to a reduced shift in the CoP at gait initiation, which is related to gait freezing [22,24]. Trunk flexion during gait is also related to gait spatiotemporal alterations [21], changes in the kinetic parameters, muscular synergies, and temporal coupling between body segments [25].

Spine kinematics during unperturbed linear gait in individuals suffering from FoG has yet not been explored. Furthermore, while the contribution of different spinal regions in determining trunk alterations has been extensively explored in PD patients with camptocormia [26] or Pisa syndrome [27], characterization of spinal lordosis and kyphosis during walking lacks adequate description and may be of great value for the design of personalized physiotherapy approaches.

In our work, we aim to describe for the first time gait spinal kinematics and their variation with respect to static balance in individuals suffering from PD and FoG. Accordingly, we re-analyzed walking and standing data previously recorded in individuals with PD and healthy controls (HCO) at our institutions [22,28]. We have proposed a simple analysis pipeline that allows for the estimation of trunk inclination and the postural asset of lower and upper spinal regions (i.e., kyphosis and lordosis angles) with the use of a limited number of markers. In addition, we investigated an unusual variable – the change in spinal length between static and dynamic posture – as it could yield insights into the overall flexibility (or lack of flexibility) of the spine.

We envisioned decreased spinal mobility and increased trunk inclination in individuals with PD suffering from FoG, with respect to both age-matched PD individuals without FoG and HCO, possibly due to increased kyphosis and reduced lordosis [29]. We also envisioned a tight relationship between spinal postural alterations and poor gait performance, as previously suggested [21,25].

With respect to previous protocols specifically designed for spinal kinematics evaluation [30–32], our method can be more easily applied in clinical practice, especially for the evaluation of individuals with PD after drug withdrawal, and are useful for retrospective analysis of already-acquired data.

## Methods

### Participants

Forty-seven volunteers with PD and 15 HCO of similar age were recruited at our laboratories. The Ethical Committees for the Milano Area 2 (Italy) and the Julius Maximilian University of Würzburg (Germany) approved the study, and all participants gave informed consent according to the Declaration of Helsinki. Exclusion criteria were previous major orthopaedic surgeries, diabetes, vestibular disorders, cardiovascular diseases, cognitive decline (Mini-Mental State Examination score ≥27), and neurological diseases other than PD. PD was diagnosed according to the United Kingdom Brain Bank Clinical Diagnostic criteria. Patients were grouped into individuals with FoG (PD-FOG, N=23) and without FoG (PD-NFOG, N=24) according to their clinical history and a clinical evaluation performed by the same neurologist with experience in movement disorders (IUI) at both centres. Severity of disease symptoms was assessed using the unified Parkinson’s disease rating scale (UPDRS) in meds-off, i.e., overnight withdrawal (>12 h) of all dopaminergic drugs, and upon receiving 1 to 1.5 times the levodopa-equivalent of the morning dose (meds-on). No patient suffered from camptocormia, Pisa syndrome, or bone deformities. Demographic and clinical data of recruited participants are shown in Table 1.

**Table 1:** Demographic, anthropometric, and clinical data. Data are shown as *median (range)*. No statistically significant differences were found across groups. Abbreviations: BMI: body mass index; HCO, healthy controls; H&Y: Hoehn and Yahr scale; LEDD: levodopa equivalent daily dose, calculated as in [63]; PD-FOG: Parkinson’s disease with freezing of gait; PD-NFOG: Parkinson’s disease with no freezing of gait; UPDRS: unified Parkinson’s disease rating scale.

|  | HCO | PD-NFOG | PD-FOG |
| --- | --- | --- | --- |
| Gender (males/total) | 8/15 | 13/23 | 16/24 |
| Age (years) | 66 (54 – 66) | 66 (45 – 79) | 64.5 (52 – 79) |
| Height (cm) | 183.3 (147 – 183.3) | 169.51 (147.22 – 187.87) | 168.33 (149.72 – 188.15) |
| Weight (Kg) | 95.6 (56 – 95.6) | 70.485 (49.15 – 108.56) | 72.78 (45.99 – 101.9) |
| BMI (kg/m<sup>2</sup>) | 32.5 (18.8 – 32.5) | 23.97 (19.71 – 34.98) | 27.47 (18.26 – 39.48) |
| Disease duration | (-) | 11 (2 – 21) | 11 (3 – 35) |
| H&Y | (-) | 2 (2 – 3) | 2 (2 – 3) |
| UPDRS-III meds-off | (-) | 27 (10 – 48) | 29 (15 – 49) |
| UPDRS-III meds-on | (-) | 9 (3 – 24) | 12 (4 – 29) |
| LEDD | (-) | 730 (180 – 1280) | 839.50 (355 – 2725) |

### Experimental setup

All patients performed the experiment after overnight suspension of all dopaminergic drugs (meds-off). Meds-off was preferred over meds-on, as most PD-FOG patients reported gait freezing during the wearing-off phases. We recorded the postural profile of each participant during a short recording (15- 60 sec) of quiet standing. Participants were asked to stand still and relax, without any voluntary movement. Participants were then instructed to walk at a comfortable pace along a linear 8 m pathway. For each subject, we collected 3-8 trials according to their compliance and clinical condition. We assessed barefoot walking as footwear modify gait characteristics in patients with PD [33]. There is also evidence of the effect of plantar stimulation on parkinsonian gait [34] and specifically on FoG [35].

### Kinematic analysis

At both centres, the same team performed data collection with the same motion capture system and marker protocol. Data were then pooled for the analysis. Pre-processing was performed by the same operators at the two centres and the analyses conducted with the same methods. Kinematics was monitored with a full-body marker set (LAMB [36–38]) and six optoelectronic cameras (SMART- E/SMART-DX, BTS, Italy; sampling frequency: 60 Hz in Italy and 100 Hz in Germany). Marker traces were filtered with a 3rd-order lowpass Butterworth filter [cut-off frequency: 10 Hz]. Spine kinematics were described by considering the spinal length, the trunk inclination, and the kyphosis and lordosis angles. To obtain these variables, the spine was modelled as composed by three linear segments (see Figure 1). Markers were placed on the seventh spinal process (C7), the maximum kyphosis point (MAX_KYPH), and the middle point between the two posterior superior iliac spines (PSIS_MX). We located the MAX_KYPH by visual inspection and palpation of the spine while participants were standing in a neutral and relaxed position, looking in front of them, and with their arms at their sides. The point was then reviewed and confirmed during voluntary forward bending of the trunk. We reconstructed the position of the coccyx based on previous radiographic studies [39,40]. Accordingly, a reference system of axes with origin in the PSIS_MX was defined. The anterior-posterior axis (x) was defined as the line connecting PSIS_MX and the middle point between the two anterior-superior iliac spines (ASIS); the vertical axis (y) was defined as the perpendicular to the anterior-posterior axis and the line connecting the right and the left ASIS; the third axis (z) was perpendicular to both the anterior-posterior and the vertical axes. From literature data, the coordinates of the coccyx in this reference frame could be estimated as [x= -0.06 m, y=-0.09 m, z=0 m] [39,40]. Accordingly, we calculated the coccyx position in the laboratory reference system by applying the following transformation:

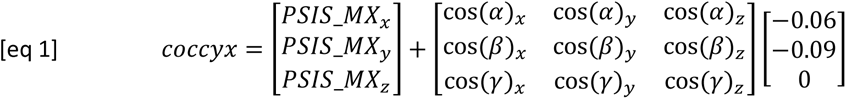

where cos(α), cos(β), and cos(γ) are the director cosines defining the orientation of the axes of the above-defined pelvis reference system.

**Figure 1:**
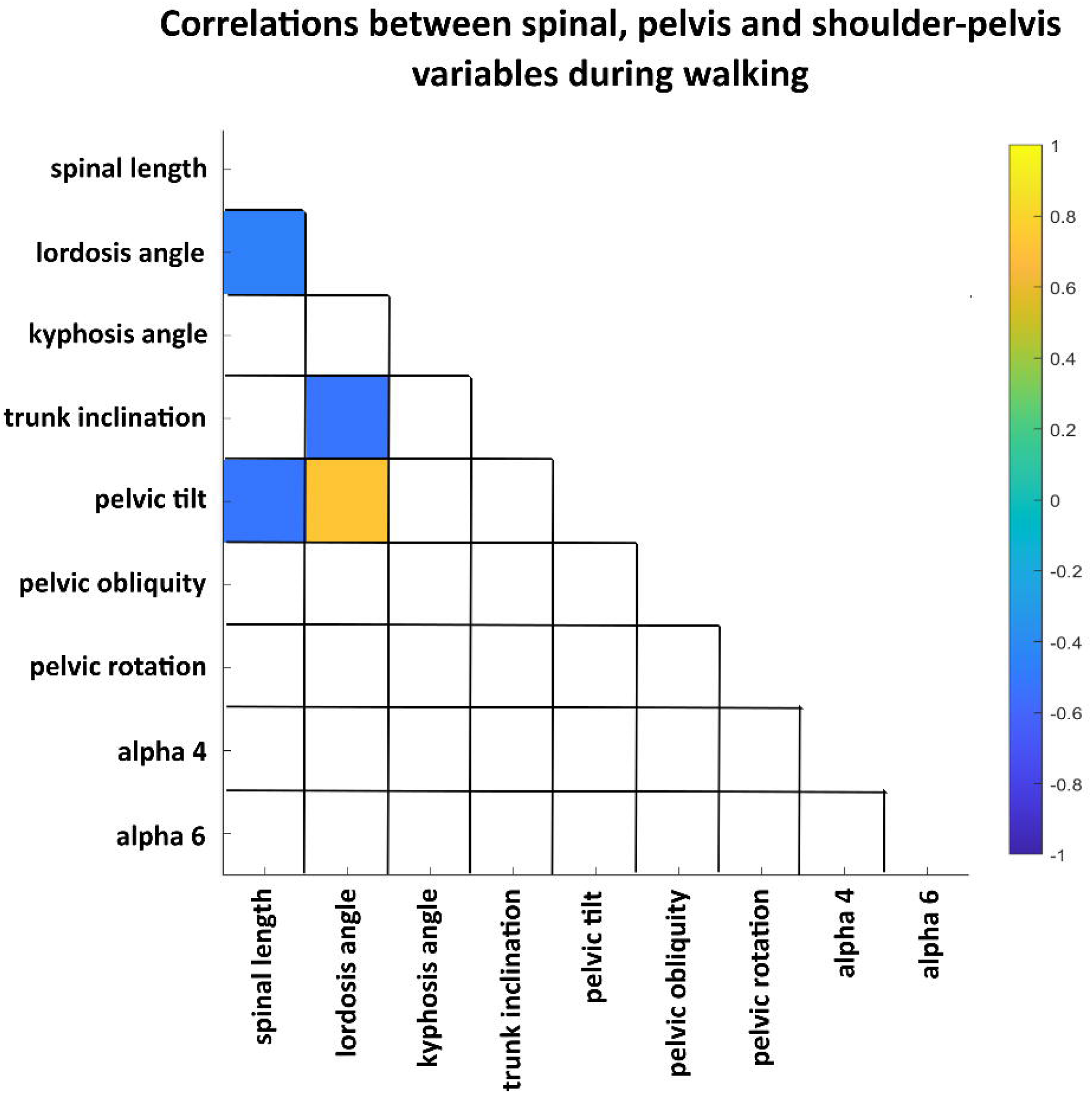
Spine variables computation. We placed three markers at the seventh cervical vertebrae (C7), the point of maximum kyphosis (MAX_KYPH), and the middle point between the posterior superior iliac spines (PSIS_MX). We then computed a virtual point for the coccyx, as a fixed translation with respect to PSIS_MX point along a reference system integral to the pelvis (eq 1). Kyphosis and lordosis angles were computed as the angles between the vectors described by these points (see the main text for details) and projected to the sagittal plane of the pelvis reference system. The spinal length (dashed line) was computed as the module of the vector between the coccyx and C7. The inclination of the vector connecting PSIS_MX and C7 with respect to the laboratory vertical axis defined trunk inclination.

We then computed the spinal length as the module of the vector connecting the markers placed on the coccyx and C7 (Figure 1) as follows:

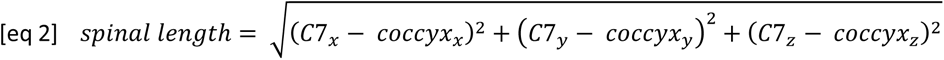

To monitor spinal length variations across conditions, we computed the percentage variation between walking (WLK) and standing (STN), as follows:

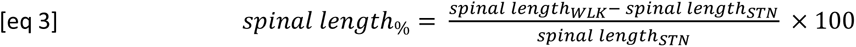

Trunk inclination was defined as the inclination of the vector connecting the markers placed on PSIS_MX and C7 with respect to the vertical axis of the laboratory (Figure 1). The kyphosis angle was computed as the angle between the vector connecting the marker on the point of maximum kyphosis (MAX_KYPH) and C7, and the vector connecting PSIS_MX and MAX_KYPH (Figure 1). The lordosis angle was estimated as the angle between the vector connecting PSIS_MX and MAX_KYPH and the coccyx and PSIS_MX (Figure 1). Both the kyphosis and lordosis angles were projected on the sagittal plane of the pelvis reference system to account for small deviations between subject orientation and laboratory axes.

The above-defined pelvis reference system allowed us to monitor the pelvis angles in the sagittal (pelvic tilt), transversal (pelvic rotation), and frontal (pelvic obliquity) laboratory planes, which have been shown to be closely related to trunk kinematics [41]. To explore the relationship between the upper trunk and pelvis, we also computed the angle between the vectors connecting the two acromion and the two ASIS in the frontal (alpha 4) and transversal (alpha 6) laboratory planes, as described in [32].

During standing, all variables were calculated and averaged along the entire acquisition time (Table 2). For walking trials, left and right heel contacts were identified based on the heel vertical trajectories, and the left and right gait cycles were defined. Only gait cycles at steady-state velocity were selected for further analyses, as described in [42]. No patient experienced gait freezing during linear walking. Spinal, pelvis, and shoulder-pelvis angles were computed during the identified gait cycles. They were then normalized in time by re-sampling their time course into one hundred points by using a cubic spline interpolation. In this way, it was possible to compute an average and a standard deviation for each variable for each data point across different gait cycles (Table 2). For each subject and variable, we also computed the average of the range across all gait cycles for the walking condition (Table 2). We computed the main spatiotemporal gait parameters by means of markers placed on the heels and halluces and averaged them across gait cycles (Table 3). All calculations were computed with ad-hoc Matlab algorithms (Matlab R2020a, The Mathworks, USA).

**Table 2:** Kinematic data for each group and condition. Data are shown as *median (min-max).* Legend: *: p<0.05, paired-sample t-test or a Wilcoxon signed rank test, as appropriate, for within-group comparisons across conditions; a,b,c,d,e: p<0.05, corrected with Dunn-Sidak or Tukey-Kramer as appropriate, for comparisons within condition across groups: a: standing HCO vs. PD-NFOG; b: standing HCO vs. PD-FOG; c: walking average values HCO vs. PD-NFOG; d: walking average values HCO vs. PD-FOG; e: walking range values HCO vs. PD-NFOG; f: walking range values HCO vs. PD-FOG; g: walking range values PDNF vs. PD-FOG. Abbreviations: HCO: healthy controls; PD-FOG: Parkinson’s disease with freezing of gait; PD-NFOG: Parkinson’s disease with no freezing of gait.

|  |  | HCO |  | PD-NFOG |  | PD-FOG |  |
| --- | --- | --- | --- | --- | --- | --- | --- |
|  |  | Standing | Walking | Standing | Walking | Standing | Walking |
| Spinal length (cm) | avg | 55.9<br>(47.8 – 59.7)<br>* | 56.5<br>(46.9 – 60.3)<br>* | 57.6<br>(45.3 – 64.3)<br>* | 57.7<br>(45.0 – 68.8)<br>* | 57.1<br>(32.7 – 65.7) | 58.3<br>(49.2 – 68.2) |
|  | %var |  | 1.2<br>(-0.3 – 2.1) |  | 0.8<br>(-1.6 – 7.8) |  | 0.4<br>(-7.7 – 7.8) |
|  | range |  | 0.7<br>(0.2 – 1.2)<br>**d** |  | 0.4<br>(0.1 – 1.7) |  | 0.3<br>(0.0 – 0.9)<br>**d** |
| Lordosis angle (°) | avg | 48.9<br>(29.1 – 60.2)<br>**a,b,\*** | 45.4<br>(30.5 – 57.0)<br>**c,d,\*** | 39.9<br>(24.6 – 51.5)<br>**a,\*** | 37.5<br>(21.9 – 50.7)<br>**c,\*** | 37.6<br>(16.1 – 51.3)<br>**b** | 36.9<br>(17.2 – 51.8)<br>**d** |
|  | range |  | 2.3<br>(0.8 – 3.7)<br>**d** |  | 1.7<br>(0.2 – 3.8) |  | 1.2<br>(0.3 – 2.9)<br>**d** |
| Kyphosis angle (°) | avg | 30.3<br>(15.7 – 37.2) | 31.2<br>(18.4 – 36.4) | 29.0<br>(15.2 – 44.4) | 28.9<br>(14.7 – 43.8) | 29.2<br>(17.6 – 45.6) | 29.2<br>(22.1 – 43.3) |
|  | range |  | 1.3<br>(0.6 – 2.8)<br>**c,d** |  | 0.9<br>(0.39 – 2.2)<br>**c** |  | 1.0<br>(0.2 – 2.3)<br>**d** |
| Trunk inclination (°) | avg | 3.9<br>(1.8 – 7.5)<br>**a,b,\*** | 7.4<br>(3.4 – 10.0)<br>**c,d,\*** | 9.3<br>(3.4 – 17.4)<br>**a,\*** | 12.1<br>(7.8 – 21.8)<br>**c,\*** | 12.7<br>(2.6 – 27.1)<br>**b,\*** | 14.3<br>(5.1 – 24.1)<br>**d,\*** |
|  | range |  | 1.9<br>(0.8 – 3.3) |  | 1.7<br>(0.7 – 3.7) |  | 1.5<br>(0.4 – 3.0) |
| Pelvic tilt (°) | avg | 8.9<br>(-10.5 – 13.4) | 9.5<br>(4.6 – 17.4) | 4.2<br>(-12.2 – 21.0) | 6.2<br>(-12.2 – 22.8) | 6.8<br>(-9.3 – 19.4) | 7.9<br>(-1.1 – 18.9) |
|  | range |  | 2.2<br>(1.1 – 3.1) |  | 2.0<br>(0.9 – 3.3) |  | 1.6<br>(0.5 – 3.5) |
| Pelvic obliquity (°) | avg | 0.4<br>(-3.9 – 2.2) | -0.4<br>(-2.2 – 1.8) | -1.0<br>(-4.0 – 2.8) | -0.5<br>(-2.3 – 2.4) | -0.2<br>(-4.0 – 3.3) | -0.3<br>(-5.5 – 4.4) |
|  | range |  | 6.7<br>(2.7 – 10.2)<br>**c,d** |  | 4.0<br>(1.5 – 8.5)<br>**c,e** |  | 3.2<br>(0.9 – 6.0)<br>**d,e** |
| Pelvic rotation (°) | avg | 0.2<br>(-4.2 – 8.0) | 2.00<br>(-3.7 – 5.6) | -0.3<br>(-6.8 – 7.0) | 0.0<br>(-4.8 – 6.2) | 1.3<br>(-4.8 – 10.0)<br>* | -0.7<br>(-4.6 – 6.1)<br>* |
|  | range |  | 7.7<br>(1.8 – 16.8) |  | 7.2<br>(3.0 – 17.0) |  | 7.2<br>(0.9 – 17.2) |
| Alpha 4 (°) | avg | -1.6<br>(-6.3 – 3.4) | -0.3<br>(-1.6 – 4.8) | -1.2<br>(-5.8 – 5.8) | -0.3<br>(-4.4 – 1.3) | -1.0<br>(-4.9 – 4.4) | 0.1<br>(-4.8 – 3.9) |
|  | range |  | 10.8<br>(6.0 – 16.7)<br>**c,d** |  | 5.3<br>(1.1 – 14.0)<br>**c** |  | 3.7<br>(0.8 – 10.2)<br>**d** |
| Alpha 6 (°) | avg | 1.8<br>(-3.1 – 5.2)<br>* | -1.1<br>(-2.7 – 1.5)<br>* | 0.6<br>(-8.2 – 6.8) | -0.2<br>(-6.0 – 3.3) | 0.1<br>(-7.2 – 7.2) | -0.1<br>(-3.1 – 9.9) |
|  | range |  | 11.5<br>(5.4 – 20.3)<br>**c,d** |  | 7.9<br>(1.9 – 17.2)<br>**c** |  | 6.6<br>(0.4 – 24.4)<br>**d** |

**Table 3:** Gait spatiotemporal parameters for the three cohorts. Data are shown as *median (min-max).* Legend: a,b: p<0.05, corrected with Dunn-Sidak or Tukey-Kramer as appropriate, a: HCO vs. PD-NFOG; b: HCO vs. PD-FOG. Abbreviations: HCO: healthy controls; PD-FOG: Parkinson’s disease with freezing of gait; PD-NFOG: Parkinson’s disease with no freezing of gait.

|  | **HCO** | **PD-NFOG** | **PD-FOG** |
| --- | --- | --- | --- |
| Cadence (step/min) | 106.16 (86.25 – 122.32) | 106.8 (75.44 – 123.29) | 105.45 (66.46 – 139.4) |
| Stride duration (s) | 1.13 (0.98 – 1.39) | 1.12 (0.97 – 1.63) | 1.15 (0.86 – 1.79) |
| Stride length (m) | 1.27 (1.03 – 1.47) **a,b** | 1.05 (0.45 – 1.39) **a** | 0.87 (0.12 – 1.26) **b** |
| Stride velocity (m/s) | 1.14 (0.74 – 1.43) **a,b** | 0.95 (0.29 – 1.34) **a** | 0.76 (0.07 – 1.31) **b** |
| Swing duration (%gait cycle) | 37.18 (30.87 – 40.2) **b** | 36.44 (26.53 – 41.51) | 35.12 (14.8 – 37.92) **b** |
| Stride width (m) | 0.089 (0.04 – 0.144) | 0.084 (0.034 – 0.172) | 0.080 (0.022 – 0.147) |

### Statistical analysis

First, we compared demographic, anthropometric, and clinical data across groups. Second, we compared gait performance by testing gait spatiotemporal parameters for differences between cohorts. Third, we tested the distribution of spinal, pelvis, and shoulder-pelvis variables for differences across conditions within each group and across groups within each condition. For the spinal length, we included in the analysis the percentage variation rather than the average value as it would have been influenced by inter-subject height differences. All other variables were analyzed in terms of average values, for standing and walking conditions, and in terms of the range of variation for the walking condition only. For each variable, group and condition, we tested the normality of the distribution with an Anderson-Darling test. Outlier values were removed by computing the jack-knifed Mahalanobis distance [43,44]. We compared demographic data across groups using a Kruskal–Wallis test or ANOVA, followed by a Dunn-Sidak or Tukey-Kramer post-hoc approach, according to data distribution. Differences in gender representation in the three cohorts were tested using a Chi-square test. Clinical data were compared between the two patient groups with a t-test or a Wilcoxon rank sum test, as appropriate. We compared conditions (i.e., standing and walking) within each group with a paired-sample t-test or a Wilcoxon signed rank test, and groups within each condition with a Kruskal–Wallis test or ANOVA, followed by Dunn-Sidak or Tukey-Kramer post-hoc approach, according to data distribution. The level of significance was set to 0.05, corrected for multiple comparisons when appropriate as described.

We then analyzed linear relationships between variables with correlation analyses. Specifically, we computed Spearman’s ρ correlation coefficients between average values of spinal, pelvis, and shoulder-pelvis variables during the walking condition, pooled across groups and within group (the level of significance was set to 0.05, Bonferroni-corrected for multiple correlations). Correlation analysis on the pooled data was intended to provide a clear representation of the kinematic chain between these variables, and supported by (i) a similar trend in the transition from standing to walking in the three groups (Table 2), (ii) the similarity between the groups in the relationship between the analysed variables (Figure S1), and (iii) the small sample size of each cohort and the limited range of variation in the variables within each group, particularly in HCO. With the same reasoning, for each gait spatiotemporal parameter significantly altered in PD cohorts, we built a simple linear regression model, each time using one different predictor among the spinal, pelvis, and shoulder-pelvis variables, on the pooled data, and in the three different cohorts. For each model, the level of significance was set to 0.05. We did not include the spinal length as predictors as it would have shown obvious relations with spatial parameters. Statistical analyses were performed with ad-hoc Matlab algorithms (Matlab R2020a, The Mathworks, USA).

## Results

Groups were matched for demographic and anthropometric measurements. Patients were also matched for clinical data (Table 1).

### Spinal, pelvis, and shoulder-pelvis variables

Spinal length increased significantly during walking with respect to standing in both HCO and PD-NFOG groups, paralleled by a decrease in the lordosis angle (Table 2). When comparing walking to standing, the trunk inclination increased in all three cohorts, the pelvic rotation decreased in the PD-FOG group only, and the alpha 6 angle decreased selectively in the HCO group (Table 2). During both standing and walking, parkinsonian individuals showed decreased lordosis angle and increased trunk inclination with respect to HCO (Table 2). During walking, the range of variation in the lordosis angle and spinal length decreased in the PD-FOG group only. The range of variations in the kyphosis angle, pelvic obliquity, alpha 4, and alpha 6 angles were affected in both patient groups, with pelvic obliquity being particularly reduced in the PD-FOG group (Table 2).

### Gait spatiotemporal parameters

Individuals with PD-NFOG and PD-FOG showed decreased stride length and velocity compared to HCO. PD-FOG additionally showed decreased swing duration compared to controls (Table 3). No differences were found between the PDNF and PDF groups.

### Correlation analyses

The analysed variables showed similar trends across groups (Figure S1), thus supporting a correlation analysis on the pooled data. The pooled data of the walking condition showed negative correlations between the spinal length and both the lordosis angle (ρ= -0.46) and the pelvic tilt (ρ= -0.51). The lordosis angle negatively correlated with trunk inclination (ρ= -0.52) and positively correlated with pelvic tilt (ρ= 0.73) (Figure 2). The only surviving correlation within groups was between the lordosis angle and pelvic tilt (HCO: ρ= 0.82; PD-NFOG: ρ= 0.86; PD-FOG: ρ= 0.76).

**Figure 2:**
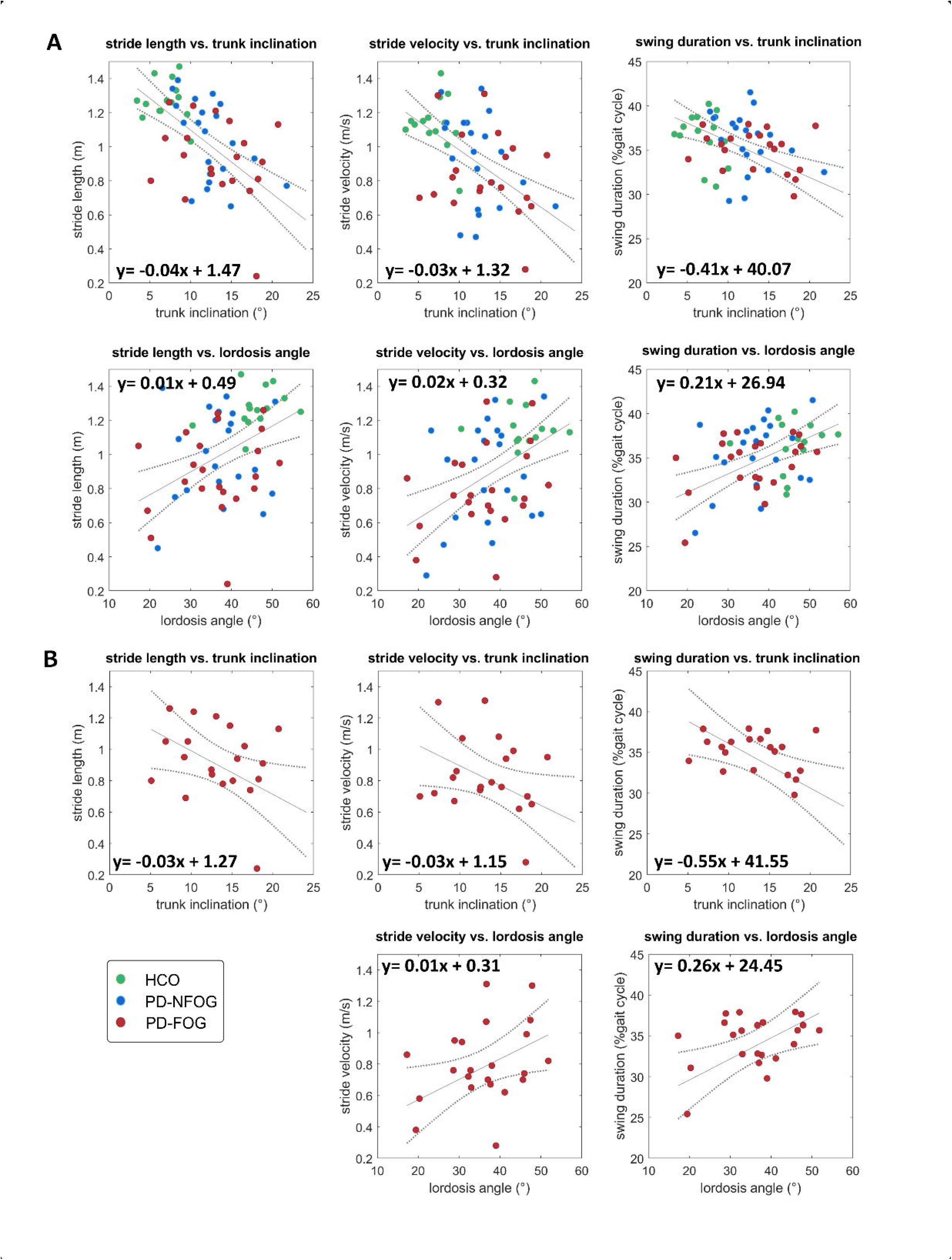
Spearman’s ρ correlation coefficient between the spinal, pelvis, and shoulder-pelvis variables during walking. As shown by the colour bar, yellow boxes indicate positive correlations while blue boxes negative correlations. Only significant results are shown (p<0.05, Bonferroni-corrected for multiple correlations).

### Regression models

When pooling the data of the three cohorts, all spatiotemporal gait parameters that showed alterations in PD groups (stride length, stride velocity, and swing duration; Table 3) were linearly related with trunk inclination and lordosis angle (Figure 3, panel A). Trunk inclination and lordosis angle showed negative and positive correlations, respectively, with stride length, stride velocity and swing duration. All other spinal, pelvis, and shoulder-pelvis variables did not yield significant results. When analyzing the three groups separately, no significant correlations existed for HCO, while only the negative correlation between trunk inclination and stride length survived in the PD-NFOG group (y= - 0.04x + 1.49). All correlations found in the pooled dataset survived in the PD-FOG group (Figure 3, panel B), apart from the correlation between the lordosis angle and stride length.

**Figure 3:**
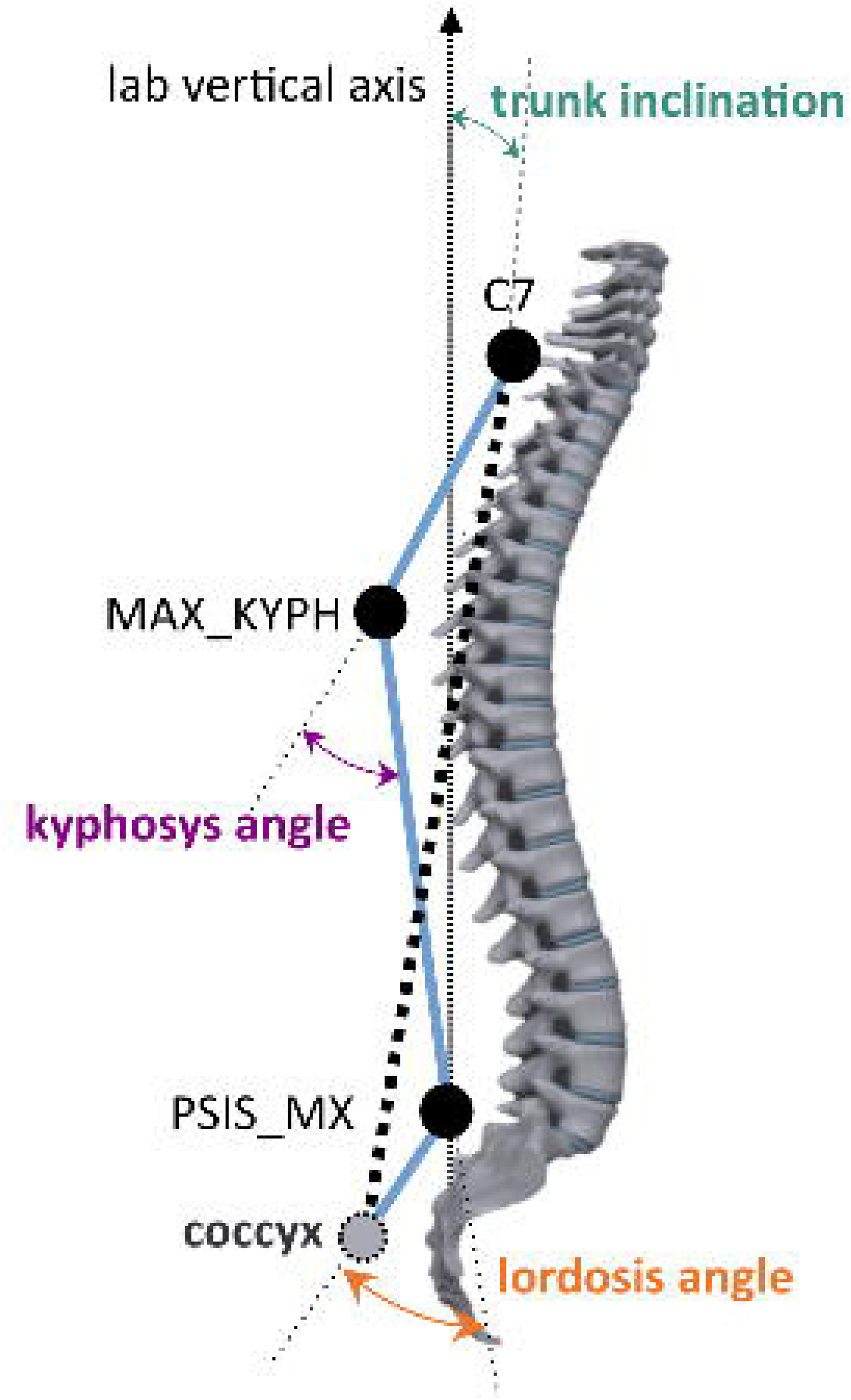
Scatter plots and linear fitting of data distribution for simple linear regression models between gait spatiotemporal parameters and spinal, pelvis, and shoulder-pelvis variables as predictors on the pooled data (panel A) and on the PD-FOG group only (panel B). Only models with significant predictors are shown (p<0.05). Only gait spatiotemporal parameters affected in PD (Table 3) were included in the analysis. Legend: green dots, HCO; blue dots: PD-NFOG; red dots: PD-FOG; solid line: fitted linear relationship; dashed line: 95% confidence bounds. Abbreviations: HCO: healthy controls; PD-FOG: Parkinson’s disease with freezing of gait; PD-NFOG: Parkinson’s disease with no freezing of gait.

## Discussion

We analyzed spine kinematics during linear walking in individuals with PD with and without FoG, to define trunk alterations peculiar to PD with FoG and to foster the development of targeted therapeutic approaches.

With respect to healthy controls, PD participants showed increased trunk inclination and lordosis angle, decreased range of variation in kyphosis angle, pelvic obliquity, and shoulder-pelvic angles, as well as characteristic gait spatiotemporal alterations, i.e. decreased stride length and velocity. Specific features of PD-FOG were poor modulation of spinal length and lordosis angle between standing and walking conditions and within walking. Of great interest, trunk inclination and lordosis angle was shown to be linearly related to altered spatiotemporal gait parameters only in the PD-FOG group.

Our results confirm previous indications of poor trunk mobility in PD [13,45–48], and show more severe impairment in those with FoG. Clinical evidence indicates a close relationship between reduced trunk mobility and FoG. Individuals with PD and FoG often experience gait freezing during turning, when the position of the CoM is controlled by the balancing of the trunk and swinging leg on the supporting hip [49]. They typically perform turnings *en-bloc*, with poor intersegmental coordination (head and trunk rotation) [50]. Overall, there seems to be a close coupling between the “passenger” and “locomotor” units in PD individuals suffering from FoG, possibly due to excessive trunk muscle tone [51] and axial rigidity [41]. This could justify the occurrence of freezing episodes when trunk action is needed to execute the motor program (e.g. during turning [49] or gait modulation [42]). During walking, excessive trunk rigidity would also prevent the attenuation of forces exchanged during locomotion and thus proper stabilization of the head, explaining the inability of fall-prone patients to cope with challenging walking conditions [16].

Considering that in our study the PD groups were matched for clinical data, axial rigidity may not be the only explanation for the poor trunk mobility in PD-FOG. Since PD individuals with FoG are more prone to falls [4], blocking of the “passenger” unit could serve as a compensatory strategy to minimize CoM perturbations in the medio-lateral direction in response to postural instability. We can also assume that individuals with PD and FoG show smaller than normal voluntary trunk displacements during walking because of poor simultaneous control of axial postural coordination and dynamic postural equilibrium in walking. Previous studies in PD have shown difficulties in coordinating multiple joints and multiple tasks [52] as possible consequence of poor proprioceptive-motor integration. In this case, our data would support a specific involvement of posterior parietal areas in individuals with PD and FoG [53], in using proprioceptive feedback to formulate internal representation of body maps for dynamic control of multisegmental movements [54–56]. The neuropathological dynamics underlying the postural and gait alterations observed in PD and FoG remain elusive and can be better clarified only using neural recordings during ongoing gait freezing episodes [57].

Our results clearly outline a deep interconnection between postural alterations and gait impairment in individuals with PD and FoG [4]. In particular, trunk inclination and lordosis angle seem to have a key role in gait performance. Specifically, trunk inclination and lordosis angle were respectively negatively and positively correlated to stride length, stride velocity, and swing duration. This relationship was observed exclusively in the PD-FOG group, and may be important for novel physical therapies. In this context, it is also important to consider that spinal variables showed deep interconnections (Figure S1 and Figure 2) as part of a single kinematic chain. From our analysis, we can infer that an increase in forward trunk inclination would relate to a decrease in the lordosis angle, as well as to increased pelvic anteversion and poor modulation of spinal length.

It is important to note that our experiment was unable to determine whether postural alterations were a determining factor or a compensatory response to gait disturbances [9]. To further explore the interconnection between posture and gait in these individuals, two possible approaches may be adopted in future studies: to observe the effect on gait performance after (i) inducing PD- and freezer- like postural changes in healthy controls or (ii) normalizing selectively postural alterations in individuals with PD and FoG. An example of the first approach can be found in the work of Jacobs and colleagues [58], who studied postural responses in healthy controls mimicking parkinsonian trunk alterations. By comparing their performance with that in PD patients, they concluded that stooped posture is destabilizing but does not account for abnormal postural responses in PD. We are not aware of studies that were able to normalize selectively postural alterations in PD patients. At the current state of knowledge, considering the interconnection between trunk and gait kinematics [4], we recommend monitoring both to ensure the best possible outcome and improve posture and gait.

Another observation from our results is the importance of assessing spinal kinematics in dynamic conditions. As previously shown [22], the PD-FOG and PD-NFOG groups showed no striking differences in the spinal asset during static posture. Distinctive features of the groups became apparent when analyzing the range of variation in kinematic variables and the relationship between spinal kinematics and gait variables under dynamic conditions. For the development of personalized physical therapies, we recommend the use of dynamic evaluations of individuals with PD and FoG to identify risk factors favouring gait freezing episodes [17,59–61].

Our study has some limitations. First, the limited number of participants may have precluded the detection of additional significant differences. Specifically, the small sample size of the HCO group may have prevented the detection of linear relationships between variables in this group. However, we are confident in our results considering that the sample size is in line with previous work on the topic, and that the PD-FOG group showed significant linear relationships despite having the highest inter-subject variability among the three cohorts. Second, computation of the lordosis angle may have underestimated the variable, as it did not rely on a marker for the maximum lordosis [32]. Our method for the calculation of the spinal variables was not confirmed by radiography investigations [31] nor a clinical assessment [62]; therefore, absolute values should be considered with caution. Still, since we have focused on the differences between groups and conditions, rather than the absolute values of the variables, we believe our approach is valid for our purposes. Further studies are warranted to explore the relationship between the spinal angles calculated with the proposed protocol and clinically defined angles [26,62], especially in the presence of camptocormia, Pisa syndrome, or bone deformities, which were not investigated in the current study and could challenge the application and repeatability of our methods. Third, we limited our evaluation to the meds-off condition. Additional studies are needed to verify spinal kinematic changes in response to therapies. Last, we did not monitor the history of falls, which could be a confounding factor for our findings. Future studies may include a group of PD with a history of falls and without FoG to disentangle changes in spine kinematics specific to falls.

In conclusion, our study highlighted the close interconnection between alterations in upper-body kinematics and gait in individuals with PD and FoG, as well as the importance of evaluating spinal changes during dynamic conditions. We showed that the main spinal alterations observed in PD individuals with a positive history of FoG are increased trunk inclination and reduced lordosis, that are directly related to shorter stride length, slower stride velocity, and longer swing time. Further studies are warranted to distinguish the nature of the observed spinal alterations, whether detrimental to walking or compensatory for walking alterations. Based on our findings, we recommend the development of physiotherapy approaches for FoG that target trunk abnormalities and gait disorders as a whole, rather than as separate entities.

## Data Availability

All data produced in the present study are available upon reasonable request to the authors

## Supplementary figures

**Figure S1:** Relationship between the most significant variables during walking within each group (HCO: green dots, PD-NFOG: blue dots, PD-FOG: red dots). The patterns were consistent across the groups, even if spanning over different ranges.

## Acknowledgements

The study was sponsored by the deutsche Forschungsgemeinschaft (DFG, German Research Foundation) – Project-ID 424778381-TRR 295, the European Union - Next Generation EU - NRRP M6C2

- Investment 2.1 Enhancement and strengthening of biomedical research in the NHS, (Project-ID PNRR- MAD-2022-12376927), the Fondazione Grigioni per il Morbo di Parkinson and the Fondazione Europea di Ricerca Biomedica (FERB Onlus). IUI was supported by a grant from New York University School of Medicine and The Marlene and Paolo Fresco Institute for Parkinson’s and Movement Disorders, which was made possible with support from Marlene and Paolo Fresco. Our special thanks go to Monica Norcini and Ilaria Riela for administrative support. Warm thanks to Dr. Elena Contaldi for her help in revising the manuscript.

## References

[1] J.G. Nutt, B.R. Bloem, N. Giladi, M. Hallett, F.B. Horak, A. Nieuwboer, Freezing of gait: Moving forward on a mysterious clinical phenomenon, Lancet Neurol. 10 (2011) 734–744. 10.1016/S1474-4422(11)70143-0.

[2] M. Michałowska, U. Fiszer, A. Krygowska-Wajs, K. Owczarek, Falls in Parkinson’s disease. Causes and impact on patients’ quality of life., Funct. Neurol. 20 (2005) 163–168.

[3] Y. Okuma, A.L. Silva de Lima, J. Fukae, B.R. Bloem, A.H. Snijders, A prospective study of falls in relation to freezing of gait and response fluctuations in Parkinson’s disease, Park. Relat. Disord. 46 (2018) 30–35. 10.1016/j.parkreldis.2017.10.013.

[4] E.M.J. Bekkers, B.W. Dijkstra, E. Heremans, S.M.P. Verschueren, B.R. Bloem, A. Nieuwboer, Balancing between the two: Are freezing of gait and postural instability in Parkinson’s disease connected?, Neurosci. Biobehav. Rev. 94 (2018) 113–125. 10.1016/j.neubiorev.2018.08.008.

[5] C. Palmisano, G. Brandt, N.G. Pozzi, L. Alice, V. Maltese, C. Andrea, V. Jens, G. Pezzoli, C.A. Frigo, I.U. Isaias, Sit-to-walk performance in Parkinson’s disease: a comparison between faller and non-faller patients, Clin. Biomech. 63 (2019) 140–146. 10.1016/j.clinbiomech.2019.03.002.

[6] R.M. Pickering, Y.A.M. Grimbergen, U. Rigney, A. Ashburn, G. Mazibrada, B. Wood, P. Gray, G. Kerr, B.R. Bloem, A meta-analysis of six prospective studies of falling in Parkinson’s disease, Mov. Disord. 22 (2007) 1892–1900. 10.1002/mds.21598.

[7] B.R. Bloem, Y.A.M. Grimbergen, M. Cramer, M. Willemsen, A.H. Zwinderman, Prospective assessment of falls in Parkinson’s disease, J. Neurol. 248 (2001) 950–958. 10.1016/S0890-5401(03)00059-2.

[8] B.R. Bloem, K.P. Bhatia, Gait and balance in basal ganglia disorders, in: A. Bronstein, T. Brandt, M. Woollacott, J. Nutt (Eds.), Clin. Disord. Balanc. Posture Gait, Arnold, London, 2004: pp. 173–206.

[9] B.R. Bloem, J.M. Hausdorff, J.E. Visser, N. Giladi, Falls and freezing of Gait in Parkinson’s disease: A review of two interconnected, episodic phenomena, Mov. Disord. 19 (2004) 871–884. 10.1002/mds.20115.

[10] O. Pelykh, A.-M. Klein, K. Bötzel, Z. Kosutzka, J. Ilmberger, Dynamics of postural control in Parkinson patients with and without symptoms of freezing of gait., Gait Posture. 42 (2015) 246–250. 10.1016/j.gaitpost.2014.09.021.

[11] G.K. Kerr, C.J. Worringham, M.H. Cole, P.F. Lacherez, J.M. Wood, P.A. Silburn, Predictors of future falls in Parkinson disease, Neurology. 75 (2010) 116–124. 10.1212/WNL.0b013e3181e7b688.

[12] M.H. Cole, P.A. Silburn, J.M. Wood, C.J. Worringham, G.K. Kerr, Falls in Parkinson’s disease: Kinematic evidence for impaired head and trunk control, Mov. Disord. 25 (2010) 2369–2378. 10.1002/mds.23292.

[13] M. Ferrarin, M. Rizzone, L. Lopiano, M. Recalcati, A. Pedotti, Effects of subthalamic nucleus stimulation and L-dopa in trunk kinematics of patients with Parkinson’s disease, Gait Posture. 19 (2004) 164–171. 10.1016/S0966-6362(03)00058-4.

[14] M. Ferrarin, M. Rizzone, B. Bergamasco, M. Lanotte, M. Recalcati, A. Pedotti, L. Lopiano, Effects of bilateral subthalamic stimulation on gait kinematics and kinetics in Parkinson’s disease, Exp. Brain Res. 160 (2005) 517–527. 10.1007/s00221-004-2036-5.

[15] G.N. Lewis, W.D. Byblow, S.E. Walt, Stride length regulation in Parkinson’s disease: The use of extrinsic, visual cues, Brain. 123 (2000) 2077–2090. 10.1093/brain/123.10.2077.

[16] M.H. Cole, G.A. Naughton, P.A. Silburn, Neuromuscular Impairments Are Associated with Impaired Head and Trunk Stability during Gait in Parkinson Fallers, Neurorehabil. Neural Repair. 31 (2017) 34–47. 10.1177/1545968316656057.

[17] M. Serrao, G. Chini, G. Caramanico, M. Bartolo, S.F. Castiglia, A. Ranavolo, C. Conte, T. Venditto, G. Coppola, C. Di Lorenzo, P. Cardinali, F. Pierelli, Prediction of responsiveness of gait variables to rehabilitation training in Parkinson’s disease, Front. Neurol. 10 (2019) 1–12. 10.3389/fneur.2019.00826.

[18] W.G. Wright, V.S. Gurfinkel, J. Nutt, F.B. Horak, P.J. Cordo, Axial hypertonicity in Parkinson’s disease: Direct measurements of trunk and hip torque, Exp Neurol. 208 (2007) 38–46. https://www.ncbi.nlm.nih.gov/pmc/articles/PMC3624763/pdf/nihms412728.pdf.

[19] R.P. Hubble, P.A. Silburn, G.A. Naughton, M.H. Cole, Trunk exercises improve balance in Parkinson disease: A phase II randomized controlled trial, J. Neurol. Phys. Ther. 43 (2019) 96–105. 10.1097/NPT.0000000000000258.

[20] J. Youn, Y. Okuma, M. Hwang, D. Kim, J.W. Cho, Falling Direction can Predict the Mechanism of Recurrent Falls in Advanced Parkinson’s Disease, Sci. Rep. 7 (2017) 3921. 10.1038/s41598-017-04302-7.

[21] D. Saha, S. Gard, S. Fatone, The effect of trunk flexion on able-bodied gait, 27 (2008) 653–660. 10.1016/j.gaitpost.2007.08.009.

[22] C. Palmisano, L. Beccaria, S. Haufe, J. Volkmann, G. Pezzoli, I.U. Isaias, Gait initiation impairment in patients with Parkinson’s disease and freezing of gait, Bioengineering. 9 (2022) 639. 10.3390/bioengineering9110639.

[23] P. Crenna, I. Carpinella, M. Rabuffetti, M. Rizzone, L. Lopiano, M. Lanotte, M. Ferrarin, Impact of subthalamic nucleus stimulation on the initiation of gait in Parkinson’s disease, Exp. Brain Res. 172 (2006) 519–532. 10.1007/s00221-006-0360-7.

[24] C. Schlenstedt, M. Muthuraman, K. Witt, B. Weisser, A. Fasano, G. Deuschl, Postural control and freezing of gait in Parkinson’s disease, Park. Relat. Disord. 24 (2016) 107–112. 10.1016/j.parkreldis.2015.12.011.

[25] R. Grasso, M. Zago, F. Lacquaniti, Interactions between posture and locomotion: Motor patterns in humans walking with bent posture versus erect posture, J. Neurophysiol. 83 (2000) 288–300. 10.1152/jn.2000.83.1.288.

[26] A. Fasano, C. Geroin, A. Berardelli, B.R. Bloem, A.J. Espay, M. Hallett, A.E. Lang, M. Tinazzi, Diagnostic criteria for camptocormia in Parkinson’s disease: a consensu-based approach, Park. Relat. Disord. (2018). 10.1016/j.parkreldis.2018.04.033.

[27] P. Barone, G. Santangelo, M. Amboni, M.T. Pellecchia, C. Vitale, Pisa syndrome in Parkinson’s disease and parkinsonism: clinical features, pathophysiology, and treatment, Lancet Neurol. 15 (2016) 1063–1074. 10.1016/S1474-4422(16)30173-9.

[28] C. Palmisano, G. Brandt, M. Vissani, N.G. Pozzi, A. Canessa, J. Brumberg, G. Marotta, J. Volkmann, A. Mazzoni, G. Pezzoli, C.A. Frigo, I.U. Isaias, Gait initiation in Parkinson’s disease: impact of dopamine depletion and initial stance condition, Front. Bioeng. Biotechnol. 8 (2020). 10.3389/fbioe.2020.00137.

[29] M. Schenkman, M. Morey, M. Kuchibhatla, Spinal flexibility and balance control among community-dwelling adults with and without Parkinson’s disease, Journals Gerontol. - Ser. A Biol. Sci. Med. Sci. 55 (2000) 441–445. 10.1093/gerona/55.8.M441.

[30] P. De Blasiis, A. Fullin, M. Sansone, A. Perna, S. Caravelli, M. Mosca, A. De Luca, A. Lucariello, Kinematic Evaluation of the Sagittal Posture during Walking in Healthy Subjects by 3D Motion Analysis Using DB- Total Protocol, J. Funct. Morphol. Kinesiol. 7 (2022). 10.3390/jfmk7030057.

[31] P. De Blasiis, A. Fullin, M. Sansone, L. Del Viscovo, F. Napolitano, C. Terracciano, G. Lus, M.A.B. Melone, S. Sampaolo, Quantitative Evaluation of Upright Posture by x-Ray and 3D Stereophotogrammetry with a New Marker Set Protocol in Late Onset Pompe Disease, J. Neuromuscul. Dis. 8 (2021) 979–988. 10.3233/JND-210663.

[32] C. Frigo, R. Carabalona, M. Dalla Mura, S. Negrini, The upper body segmental movements during walking by young females, Clin. Biomech. 18 (2003) 419–425. 10.1016/S0268-0033(03)00028-7.

[33] M.A. Horn, A. Gulberti, U. Hidding, C. Gerloff, W. Hamel, C.K.E. Moll, M. Pötter-Nerger, Comparison of Shod and Unshod Gait in Patients With Parkinson’s Disease With Subthalamic and Nigral Stimulation, Front. Hum. Neurosci. 15 (2022). 10.3389/fnhum.2021.751242.

[34] L. Brognara, O. Cauli, Mechanical Plantar Foot Stimulation in Parkinson′s Disease: A Scoping Review, Diseases. 8 (2020) 12. 10.3390/diseases8020012.

[35] W. Phuenpathom, P. Panyakaew, P. Vateekul, D. Surangsrirat, A. Hiransuthikul, R. Bhidayasiri, Vibratory and plantar pressure stimulation: Steps to improve freezing of gait in Parkinson’s disease, Parkinsonism Relat. Disord. 105 (2022) 43–51. 10.1016/j.parkreldis.2022.10.024.

[36] V. Farinelli, C. Palmisano, S.M. Marchese, C.M.M. Strano, S. D’Arrigo, C. Pantaleoni, A. Ardissone, N. Nardocci, R. Esposti, P. Cavallari, Postural control in children with cerebellar ataxia, Appl. Sci. 10 (2020) 1–13. 10.3390/app10051606.

[37] A. Ferrari, M.G. Benedetti, E. Pavan, C. Frigo, D. Bettinelli, M. Rabuffetti, P. Crenna, A. Leardini, Quantitative comparison of five current protocols in gait analysis, Gait Posture. 28 (2008) 207–216. 10.1016/j.gaitpost.2007.11.009.

[38] C. Palmisano, M. Todisco, G. Marotta, J. Volkmann, C. Pacchetti, C.A. Frigo, G. Pezzoli, I.U. Isaias, Gait initiation in progressive supranuclear palsy : brain metabolic correlates, NeuroImage Clin. 28 (2020) 102408. 10.1016/j.nicl.2020.102408.

[39] A. Tsuruta, J. Tashiro, T. Ishii, Y. Oka, A. Suzuki, H. Kondo, S. Yamaguchi, Prediction of anastomotic leakage after laparoscopic low anterior resection in male rectal cancer by pelvic measurement in magnetic resonance imaging, Surg. Laparosc. Endosc. Percutaneous Tech. 27 (2017) 54–59. 10.1097/SLE.0000000000000366.

[40] J.T.K. Woon, V. Perumal, J.Y. Maigne, M.D. Stringer, CT morphology and morphometry of the normal adult coccyx, Eur. Spine J. 22 (2013) 863–870. 10.1007/s00586-012-2595-2.

[41] R.E.A. Van Emmerik, R.C. Wagenaar, A. Winogrodzka, E.C. Wolters, Identification of axial rigidity during locomotion in parkinson disease, Arch. Phys. Med. Rehabil. 80 (1999) 186–191. 10.1016/S0003-9993(99)90119-3.

[42] C. Palmisano, P. Kullmann, I. Hanafi, M. Verrecchia, M.E. Latoschik, A. Canessa, M. Fischbach, I.U. Isaias, A Fully-Immersive Virtual Reality Setup to Study Gait Modulation, Front. Hum. Neurosci. 16 (2022) 783452. 10.3389/fnhum.2022.783452 Objective:

[43] V. Farinelli, L. Hosseinzadeh, C. Palmisano, C. Frigo, An easily applicable method to analyse the ankle-foot power absorption and production during walking, Gait Posture. 71 (2019) 56–61. 10.1016/j.gaitpost.2019.04.010.

[44] K.I. Penny, Appropriate Critical Values When Testing for a Single Multivariate Outlier by Using the Mahalanobis Distance, J. R. Stat. Soc. Ser. C (Applied Stat. 45 (1996) 73–81. 10.2307/2986224.

[45] K.J. Bridgewater, M.H. Sharpe, Trunk muscle performance in early Parkinson’s disease, Phys. Ther. 78 (1998) 566–576. 10.1093/ptj/78.6.566.

[46] E. Nikfekr, K. Kerr, S. Attfield, D.E. Playford, Trunk movement in Parkinson’s disease during rising from seated position, Mov. Disord. 17 (2002) 274–282. 10.1002/mds.10073.

[47] M. Rizzone, M. Ferrarin, A. Pedotti, B. Bergamasco, E. Bosticco, M. Lanotte, P. Perozzo, A. Tavella, E. Torre, M. Recalcati, A. Melcarne, L. Lopiano, High-frequency electrical stimulation of the subthalamic nucleus in Parkinson’s disease: Kinetic and kinematic gait analysis, Neurol. Sci. 23 (2002) 103–104. 10.1007/s100720200090.

[48] T. Mitchell, D. Conradsson, C. Paquette, Gait and trunk kinematics during prolonged turning in Parkinson’s disease with freezing of gait, Park. Relat. Disord. 64 (2019) 188–193. 10.1016/j.parkreldis.2019.04.011.

[49] K. Hase, R.B. Stein, Turning strategies during human walking, J. Neurophysiol. 81 (1999) 2914–2922. 10.1152/jn.1999.81.6.2914.

[50] G. Verheyden, A.M. Willems, L. Ooms, A. Nieuwboer, Validity of the Trunk Impairment Scale as a Measure of Trunk Performance in People With Parkinson’s Disease, Arch. Phys. Med. Rehabil. 88 (2007) 1304– 1308. 10.1016/j.apmr.2007.06.772.

[51] W.G. Wright, V.S. Gurfinkel, J. Nutt, F.B. Horak, P.J. Cordo, Axial hypertonicity in Parkinson’s disease: Direct measurements of trunk and hip torque, Exp. Neurol. 208 (2007) 38–46. 10.1016/j.expneurol.2007.07.002.

[52] H. Poizner, O.I. Fookson, M.B. Berkinblit, W. Hening, G. Feldman, S. Adamovich, Pointing to remembered targets in 3-D space in Parkinson’s disease., Motor Control. 2 (1998) 251–277. 10.1123/mcj.2.3.251.

[53] C. Tard, A. Delval, D. Devos, R. Lopes, P. Lenfant, K. Dujardin, C. Hossein-Foucher, F. Semah, A. Duhamel, L. Defebvre, F. Le Jeune, C. Moreau, Brain metabolic abnormalities during gait with freezing in Parkinson’s disease, Neuroscience. 307 (2015) 281–301. 10.1016/j.neuroscience.2015.08.063.

[54] J. V. Jacobs, F.B. Horak, Abnormal proprioceptive-motor integration contributes to hypometric postural responses of subjects with parkinson’s disease, Neuroscience. 141 (2006) 999–1009. 10.1016/j.neuroscience.2006.04.014.

[55] M. Tagliabue, G. Ferrigno, F. Horak, Effects of Parkinson’s disease on proprioceptive control of posture and reaching while standing, Neuroscience. 158 (2009) 1206–1214. 10.1016/j.neuroscience.2008.12.007.

[56] M. Vaugoyeau, H. Hakam, J.P. Azulay, Proprioceptive impairment and postural orientation control in Parkinson’s disease, Hum. Mov. Sci. 30 (2011) 405–414. 10.1016/j.humov.2010.10.006.

[57] N.G. Pozzi, A. Canessa, C. Palmisano, J. Brumberg, F. Steigerwald, M.M. Reich, B. Minafra, C. Pacchetti, G. Pezzoli, J. Volkmann, I.U. Isaias, Freezing of gait in Parkinson’s disease reflects a sudden derangement of locomotor network dynamics, Brain. 142 (2019) 2037–2050. 10.1093/brain/awz141.

[58] J. V. Jacobs, D.M. Dimitrova, J.G. Nutt, F.B. Horak, Can stooped posture explain multidirectional postural instability in patients with Parkinson’s disease?, Exp. Brain Res. 166 (2005) 78–88. 10.1007/s00221-005-2346-2.

[59] M. Gandolfi, M. Tinazzi, F. Magrinelli, G. Busselli, E. Dimitrova, N. Polo, P. Manganotti, A. Fasano, N. Smania, C. Geroin, Four-week trunk-specific exercise program decreases forward trunk flexion in Parkinson’s disease: A single-blinded, randomized controlled trial, Park. Relat. Disord. 64 (2019) 268–274. 10.1016/j.parkreldis.2019.05.006.

[60] D. Volpe, M.G. Giantin, P. Manuela, C. Filippetto, E. Pelosin, G. Abbruzzese, A. Antonini, Water-based vs. non-water-based physiotherapy for rehabilitation of postural deformities in Parkinson’s disease: a randomized controlled pilot study, Clin. Rehabil. 31 (2017) 1107–1115. 10.1177/0269215516664122.

[61] M. Bartolo, M. Serrao, C. Tassorelli, R. Don, A. Ranavolo, F. Draicchio, C. Pacchetti, S. Buscone, A. Perrotta, A. Furnari, P. Bramanti, L. Padua, F. Pierelli, G. Sandrini, Four-week trunk-specific rehabilitation treatment improves lateral trunk flexion in Parkinson’s disease, Mov. Disord. 25 (2010) 325–331. 10.1002/mds.23007.

[62] N.G. Margraf, R. Wolke, O. Granert, A. Berardelli, B.R. Bloem, R. Djaldetti, A.J. Espay, A. Fasano, Y. Furusawa, N. Giladi, M. Hallett, J. Jankovic, M. Murata, M. Tinazzi, J. Volkmann, D. Berg, G. Deuschl, Consensus for the measurement of the camptocormia angle in the standing patient., Parkinsonism Relat. Disord. 52 (2018) 1–5. 10.1016/j.parkreldis.2018.06.013.

[63] C.L. Tomlinson, R. Stowe, S. Patel, C. Rick, R. Gray, C.E. Clarke, Systematic review of levodopa dose equivalency reporting in Parkinson’s disease, Mov. Disord. 25 (2010) 2649–2653. 10.1002/mds.23429.

