## Supplementary figure 1 for "Dynamic evaluation of spine kinematics in individuals with Parkinson’s disease and freezing of gait"

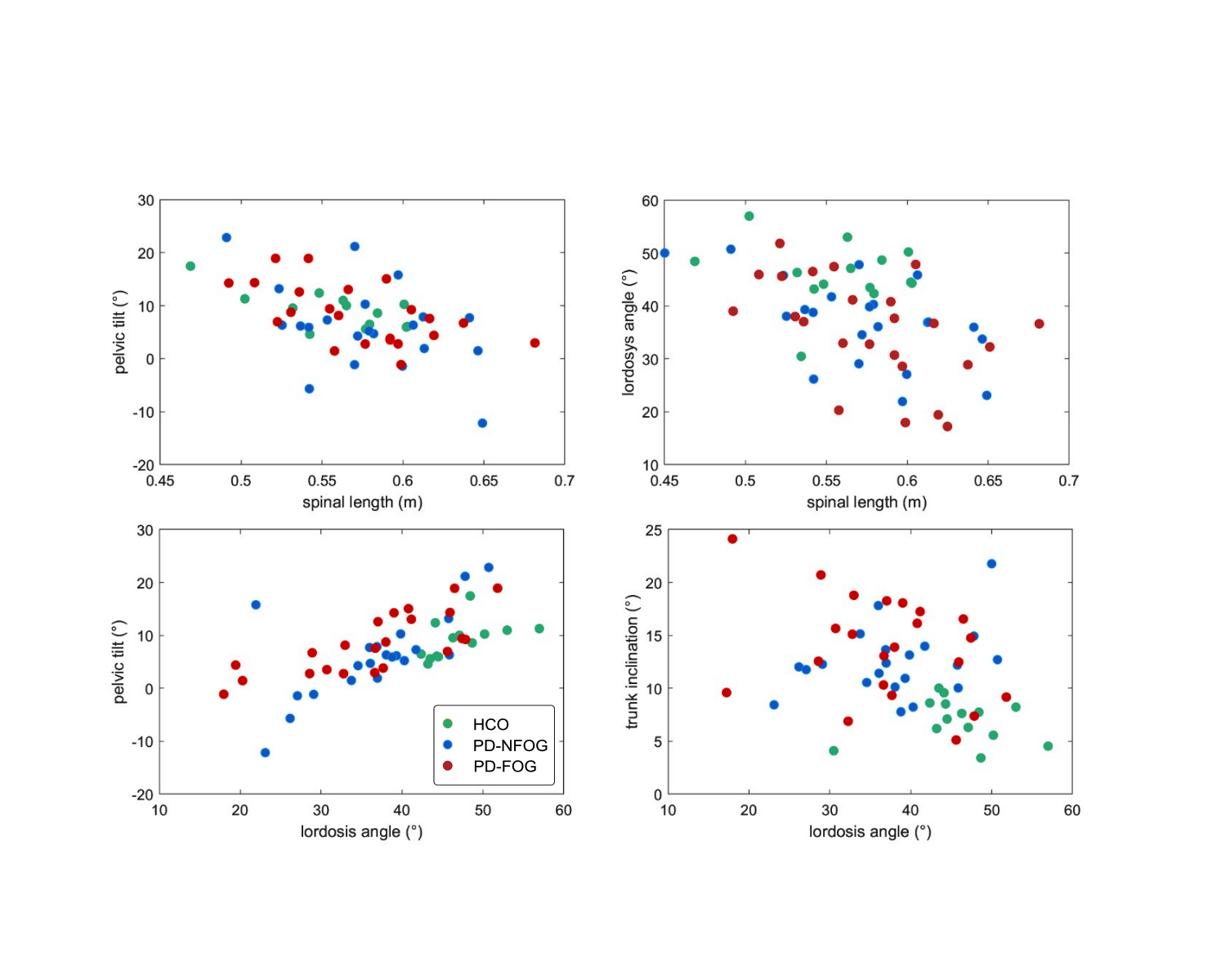

**Figure S1**: Relationship between the most significant variables during walking within each group (HCO: green dots, PD-NFOG: blue dots, PD-FOG: red dots). The patterns were consistent across the groups, even if spanning over different ranges.
